# Metastatic breast cancer patient perception of somatic tumor genomic testing

**DOI:** 10.1101/2020.01.20.19015404

**Authors:** Elizabeth J. Adams, Sarah Asad, Raquel Reinbolt, Katharine A. Collier, Mahmoud Abdel-Rasoul, Susan Gillespie, James L. Chen, Mathew A. Cherian, Anne M. Noonan, Sagar Sardesai, Jeffrey VanDeusen, Robert Wesolowski, Nicole Williams, Charles L. Shapiro, Erin R. Macrae, Robert Pilarski, Amanda E. Toland, Leigha Senter, Bhuvaneswari Ramaswamy, Clara N. Lee, Maryam B. Lustberg, Daniel G. Stover

## Abstract

**Purpose:** To assess metastatic breast cancer (MBC) patient perceptions and comprehension of tumor genomic testing and to evaluate associations with psychological wellbeing.

**Methods:** In a prospective, single institution, single-arm trial, patients with MBC underwent next-generation sequencing at study entry, with sequencing results released at progression. Patients who completed surveys before undergoing sequencing were included in the study (n=58). We administered four validated psychosocial measures: Center for Epidemiologic Studies Depression Scale, Beck Anxiety Inventory, Trust in Physician Scale, and Communication and Attitudinal Self-Efficacy scale for Cancer. Genetic comprehension was assessed using 7-question objective and 6-question subjective measures. Longitudinal data were assessed using paired Wilcoxon signed rank and McNemar’s test of agreement.

**Results:** There were no significant differences between the beginning and end of study in depression, anxiety, physician trust, or self-efficacy (median time on study: 7.6 months). Depression and anxiety were positively associated with each other and both negatively associated with self-efficacy. Self-efficacy decreased from pre-to post-genomic testing (p=0.05). There was a wide range of objective genetics comprehension and comprehension was significantly lower in non-white patients (p=0.02) and patients with lower income (p=0.04). Patients expressed increased confidence in their ability to teach others about genetics at end of study.

**Conclusions:** This is the only study, to our knowledge, to longitudinally evaluate multiple psychological metrics in MBC as patients undergo tumor genomic testing. Among patients with MBC, depression and anxiety metrics were negatively correlated with patient self-efficacy. Patients undergoing somatic genomic testing had limited genomic knowledge, which varied by demographic groups.

## INTRODUCTION

More than 150,000 women are living with metastatic breast cancer (MBC) in the United States alone.^1^ Because of increasing evidence that specific somatic tumor mutations are both prognostic (e.g. *ESR1*^2-4^) and predictive in MBC (e.g. *ESR1*^3^ and *PIK3CA*^4-6^), somatic genomic testing has become standard of care and offers great promise in the advancement of novel therapeutics and precision cancer medicine.^7-10^ Despite the richness of information next-generation sequencing (NGS) offers clinicians, the effects of genomic testing, particularly somatic tumor sequencing, on patient psychosocial outcomes and perceptions of care is understudied.^11-14^ Genetic testing may increase negative emotions in the metastatic cancer population,^14^ which in turn may decrease quality of life.^15-18^ With recent widespread implementation of tumor genomic testing, it is imperative to understand patient understanding and perceptions of genomic testing.^13^

We previously completed a prospective, single institution, single-arm trial, in which patients with MBC underwent NGS using Foundation Medicine to evaluate somatic cell mutations associated with 315 cancer-related genes^19^ at study entry, with sequencing results released to providers at time of progression.^20^ We previously found that NGS impacted clinical decision-making in a minority of patients, and patients whose next cancer treatment was not supported by the genomic test had poorer perceptions of their care.^20^ While several studies have reported the nuanced psychosocial effects of learning of one’s harbored mutations from genetic testing, most have focused on germline sequencing,^21-23^ with a relative paucity of data regarding somatic genomic testing.

We sought to evaluate patient psychosocial characteristics, genetic comprehension, and perceived risks and expectations of somatic cell NGS genomic testing in the MBC setting in this secondary analysis. The primary objectives of this study were to 1) longitudinally describe patient psychological health before and after undergoing tumor NGS; 2) evaluate the relationship between psychological health and whether the patient underwent a FM-supported treatment; 3) assess patient comprehension of genetics and association with patient perceptions of care; 4) describe the relationship between patient psychosocial outcomes and sociocultural background.

## METHODS

### Study Population

This was a prospective, single-site, single-arm trial at an NCI-designated comprehensive cancer center. Patients with MBC who were within 10 weeks of starting their current line of therapy and had an estimated survival of ≥3 months were included in this study. Participants also needed to have a tumor sample (primary or metastatic) available for genomic testing. The study population along with a description of the genomic testing has been previously described.^20^ In the present manuscript, the study population includes participants who completed the pre-test (prior to genomic testing) questionnaire at study entry (n=58) and/or the post-test questionnaire at the end of study visit (n=40). All patients completed informed consent approved by the Ohio State University Institutional Review Board, which included a description of the somatic genomic testing and discussion of study risks and benefits with their treating physician. Genomic test results were reviewed with participants; there was not a specific script for interactions or education provided beyond standard provider discussion.

### Survey Measures

Patient surveys included questions about mood, attitudes, and knowledge relating to cancer care and genomic testing. Four validated measures were used, including: Center for Epidemiologic Studies Depression Scale (CES-D),^24^ Beck Anxiety Inventory (BAI),^25^ Trust in Physicians/Providers Scale (TPS),^26^ and Communication and Attitudinal Self-Efficacy scale for cancer (CASE-cancer).^27^ The CES-D utilizes 20 items to evaluate clinical depression^24^ to determine the prevalence of depressive ideologies in the participant’s preceding week. Response options to each item are 0 (rarely or none of the time), 1 (some or little of the time), 2 (moderately or much of the time), or 3 (most or all of the time). CES-D scores range from 0 to 60 with a score of 16 or higher indicating clinical depression.^24^ The 21-question Beck Anxiety Inventory (BAI)^25^ assesses the extent of bothersome anxiety symptoms in the past month using a 4-point Likert scale. Response options to each item are 0 (not at all bothered), 1 (mildly bothered), 2 (moderately bothered), and 3 (severely bothered). BAI scores range from 0 to 63 with a score from 0-9 indicating no anxiety, 10-18 indicating mild to moderate anxiety, and a score of 19 or higher indicating moderate to severe anxiety.^25^

The 11-item Trust in Physician Survey (TPS)^26^ uses a 5-point Likert scale that includes responses of agreement ranging from 0 (strongly disagree) to 4 (strongly agree). TPS scores range from 11 to 55 with higher scores indicating greater trust in physician.^26^ The Communication and Self-Efficacy scale for cancer (CASE-cancer) evaluates patients’ confidence in maneuvering through their cancer care and relationship with their medical oncologist.^27^ CASE-cancer utilizes a 5-point Likert scale ranging from 1 (strongly disagree) to 5 (strongly agree) to assess agreement with 12 items relating to patient self-efficacy. CASE-cancer scores range from 12 to 48 with higher scores indicating greater self-efficacy.^27^ Patients who skipped more than two questions on a given measure did not have a score calculated.

Subjective and objective genetic knowledge were measured with items developed for this study. The response options for the 6 subjective questions were a 5-point Likert scale from strongly disagree to strongly agree. Objective knowledge was measured with 15 true or false statements designed to test patients’ objective understanding of genes and genetics. We selected the seven statements about basic genetic information to assess. Examples of these statements include “It is possible to see a gene with the naked eye”, “A gene is a piece of DNA”, and “A person’s race and ethnicity can affect how likely they are to get a disease”.

Physicians received a 5-item questionnaire after the FM report was released to assess whether or not they changed their treatment recommendation based on these results. The physician questionnaire has been previously described.^20^

## Statistical Analyses

One-way ANOVA tests were used to assess pre-test CES-D, BAI, TPS, and CASE-Cancer scores by income, education, insurance, and breast cancer receptor subtype. An independent t-test was used to evaluate each patient reported outcome by race (white versus non-white). Paired Wilcoxon signed-rank tests were used to compare pre- and post-test scores. McNemar’s test of agreement was used to compare pre- and post-test subjective knowledge about genetics. Due to our limited sample size, these questions were assessed as a 3-level outcome (agree vs. neutral vs. disagree). The objective knowledge questions were scored like a test (“gene test” score= number of statements answered correctly/7) and combined into a continuous variable. Linear mixed effects models were fit to assess the association between treatment change and post-test scores for each of the four outcomes. These models included main effects of treatment change (no change versus change), time (pre vs. post), the interaction of treatment change with time, and random intercepts to account for repeated measures. Finally, Pearson’s r correlation tests were performed to assess the associations among the outcomes. All figures were created in R version 3.4.1. All statistical analyses were performed using SAS version 9.4 and R version 3.4.1.

## RESULTS

### Study Population

As described previously,^20^ a total of 100 patients had successful FM NGS testing and were eligible for analysis (**Figure 1**). Of these, 58 patients completed at least a portion of the pre-test, and 40 completed at least a portion of both the pre-test and post-test surveys. Eleven patients who completed the pre-test survey did not have their FM reports released and thus did not complete a post-test survey because they did not progress on their first treatment. We assessed for potential response bias and found that patients who completed the pre-test only did not differ from those who completed both surveys in terms of demographics such as age, race, breast cancer type, or ECOG status. The 58 patients who completed the pre-test survey were mostly aged 45-64 (65%), white (81%), ER+ (61%), in the highest income (59%) group, and had private insurance (73%) (**Table 1**).

**Table 1.** Cohort Characteristics (N=58)

|  | **N** | **%** |
| --- | --- | --- |
| **Age** |  |  |
| <45 | 7 | 12.1 |
| 45-54 | 19 | 32.8 |
| 55-64 | 17 | 29.3 |
| ≥65 | 15 | 25.9 |
| **Subtype** |  |  |
| ER and/or PR+, HER2- | 35 | 61.4 |
| HER2+ | 3 | 5.3 |
| TNBC | 19 | 33.3 |
| **Race** |  |  |
| Non-white | 11 | 19.0 |
| White | 47 | 81.0 |
| **Education** |  |  |
| High school diploma/GED or lower | 17 | 29.3 |
| Some college, technical school, or Associate's degree | 20 | 34.5 |
| Bachelor's degree or higher | 21 | 36.2 |
| **Income** |  |  |
| <\$40,000 | 10 | 21.7 |
| \$40,000-\$69,999 | 9 | 19.6 |
| \$70,000 or more | 27 | 58.7 |
| **Insurance type** |  |  |
| Medicaid or Medicare | 12 | 21.4 |
| Private | 41 | 73.2 |
| Other | 3 | 5.4 |
| **Treatment change based on FM test** |  |  |
| No | 41 | 87.2 |
| Yes | 6 | 12.8 |

**Figure 1.**
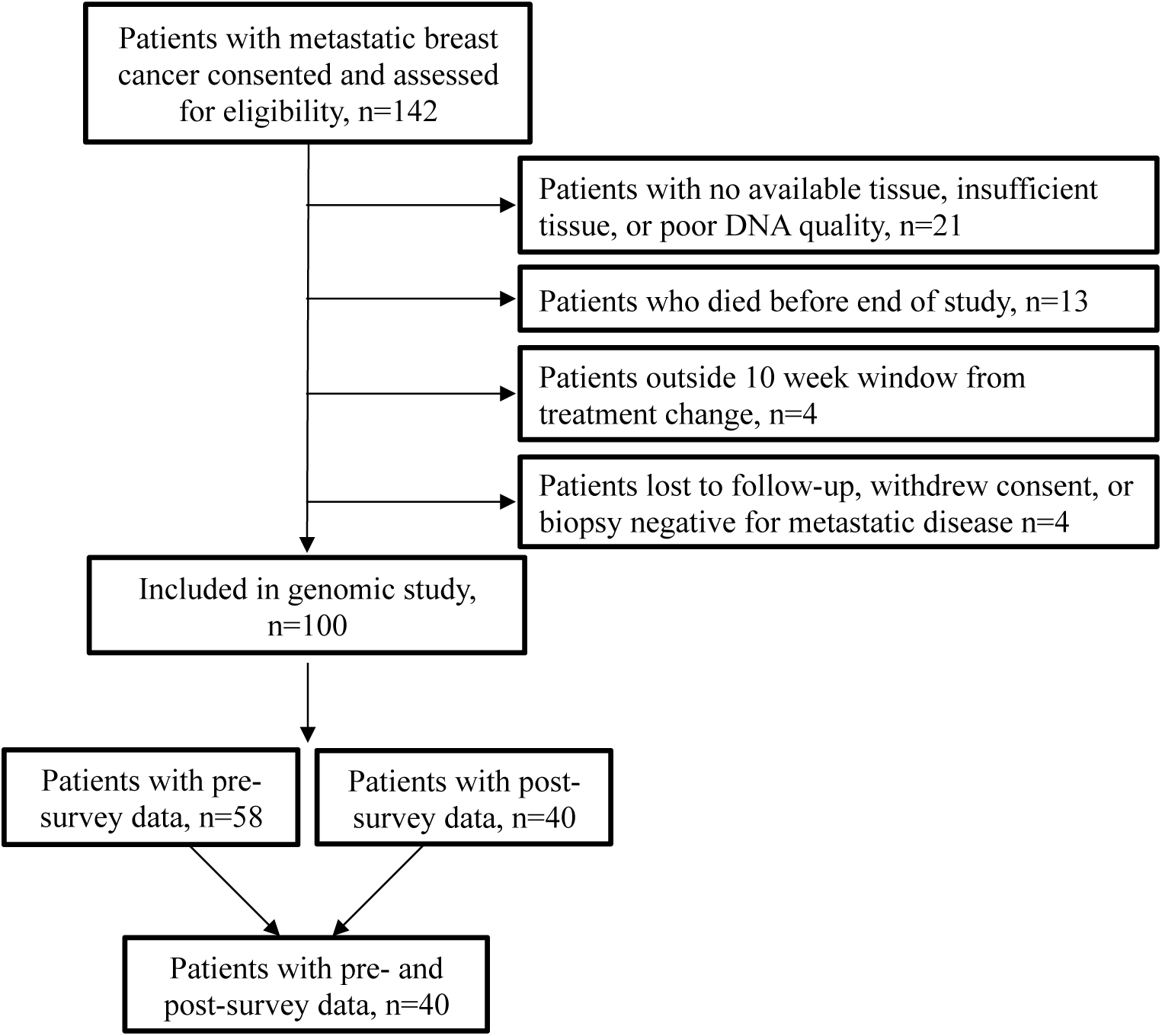
CONSORT Diagram.

### Depression, Anxiety, Physician Trust, and Self-Efficacy Measures

In the pre-test, patients had a mean CES-D score of 13.6 (range: 0-34) and a mean BAI score of 11.3 (range: 0-40). Categorically, 38% of patients were depressed (CES-D≥16), 28% of patients had mild to moderate anxiety (BAI 10-18), and 19% of patients had moderate to severe anxiety (BAI ≥19). Patients had a mean TPS score of 48.4 (range: 34-55), indicating substantial trust in their medical oncologist. Patients had a mean CASE score of 42.4 (range: 16-48), indicating relatively high self-efficacy in navigation of their cancer care. There were no significant differences in any of the outcomes by income (all ANOVA p>0.05), education (all ANOVA p>0.05), race (all t-test p>0.05), insurance (all ANOVA p>0.05), or breast cancer type (all ANOVA p>0.05) (**Supplementary Figure 1**).

From pre-test (study entry) to post-test (end of study), there was no significant change in CES-D score (Wilcoxon p-value= 0.13), BAI score (Wilcoxon p-value= 0.50), TPS score (Wilcoxon p-value= 0.15), while CASE score decreased from pre-to post-test, marginally non-significant (Wilcoxon p-value= 0.05) (**Figure 2A**). Finally, treatment change was not a significant predictor of post-test CES-D score, BAI score, TPS score, or CASE score in linear mixed model analyses (**Supplementary Table 1)**.

**Figure 2.**
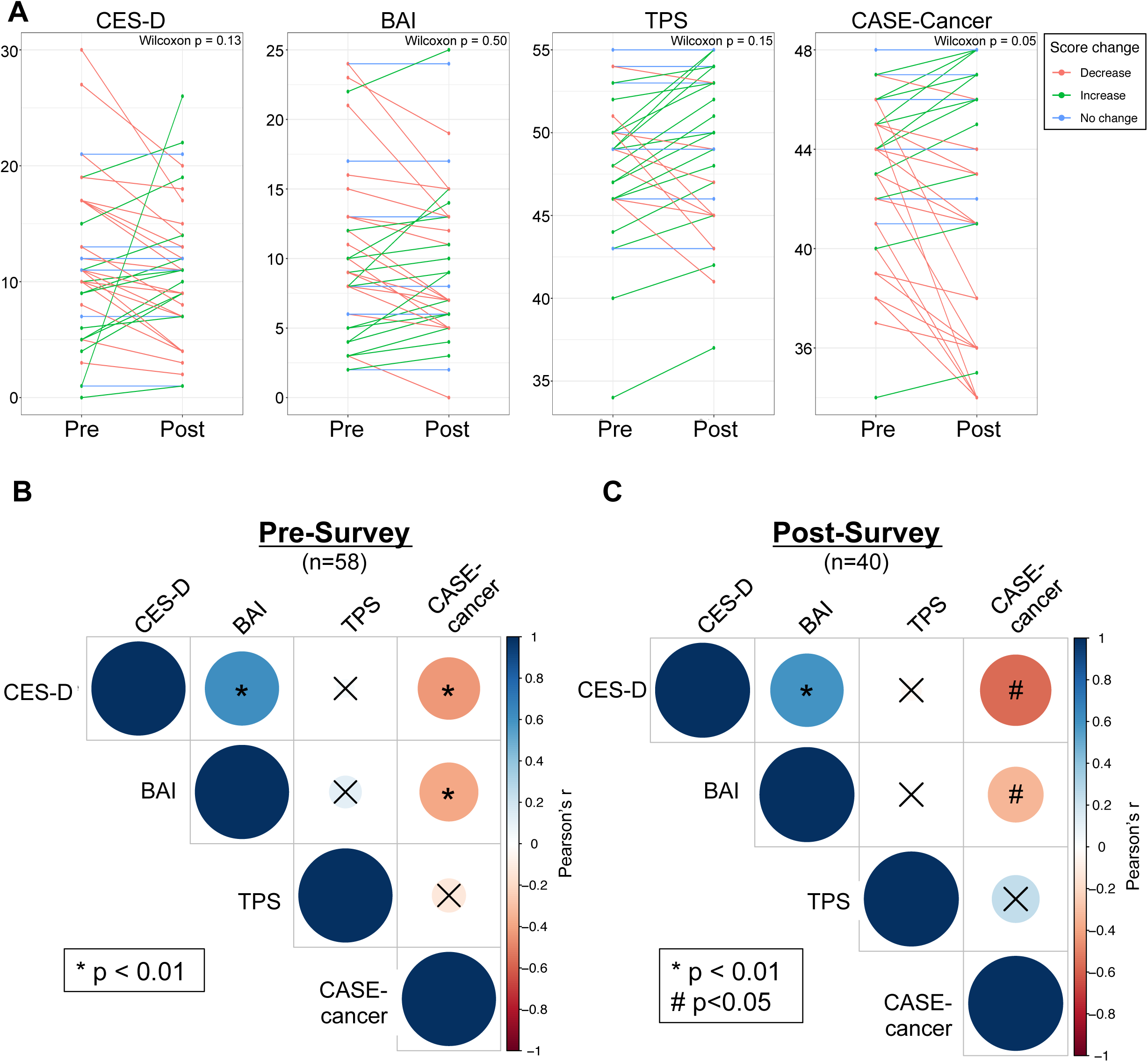
Assessment and Change in Validated Psychological Metrics. Four validated psychosocial measures were assessed in patients at study entry: Center for Epidemiologic Studies Depression Scale (CES-D)^24^, Beck Anxiety Inventory (BAI) ^25^, Trust in Physicians/Providers Scale (TPS)^26^, and Communication and Attitudinal Self-Efficacy scale for cancer (CASE-cancer)^27^. **A**. Change in each measure was evaluated for those patients who completed both ‘pre’ and ‘post’ assessments (n=40 patients). Direction of change in score is indicated in color as decrease (red), increase (green), and no change (blue). Association was assessed by Wilcoxon signed rank test. **B/C**. Correlation between each validated metric and all other metrics was assessed for all patients at study entry (**B;** n=58) and at end of study (**C**; n=40). Direction correlation (Pearson’s r) is indicated by the color of each dot (positive correlation in blue, negative correlation in red) and magnitude of correlation indicated by size of each dot (higher correlation is larger size). Associations that were not statistically significant are indicated with a black ‘x.’

We assessed correlations among our primary outcomes including CES-D, BAI, CASE, and TPS scores (**Figure 2B/C**). CES-D score was positively correlated with BAI score (Pearson’s r=0.61 and 0.60 for pre-test and post-test, respectively; both p<0.0001). CASE score was negatively correlated with CES-D and BAI in both the pre-test (Pearson’s r=-0.43 and -0.42 for CES-D and BAI pre-test, respectively; both p=0.001) and the post-test (Pearson’s r=-0.54 and -0.37 for CES-D and BAI post-test; p= 0.0004 and 0.03, respectively).

### Subjective and Objective Genetic Knowledge

Six subjective genetic knowledge questions were compared between pre-test and post-test (**Table 2**). For questions 1-5, most patients agreed with the statements indicating high confidence in understanding genetic information. For these five questions, there were no significant changes from pre-to post-test. However, for question 6 about ability to explain to others how genes affect health, patients were equally spread across response values in the pre-test. For this question, there was a significant change from pre-to post-test (McNemars p-value= 0.04). In the post-test, significantly more patients (18 vs. 13 in the pre-test) agreed that they would be able to explain to others how genes affect health.

**Table 2.** Subjective Knowledge about Genetics

|  |  |  |  |  |
| --- | --- | --- | --- | --- |
| **1. You are confident in your ability to understand information about genetics.** |  |  |  |  |
| **Pre-test N (%)** | **Post-test N (%)** |  |  | **McNemar's test p-value\*** |
|  | Disagree | Neutral | Agree | 0.19 |
| Disagree | 4 (10.3) | 3 (7.7) | 0 (0.0) |  |
| Neutral | 0 (0.0) | 5 (12.8) | 4 (10.3) |  |
| Agree | 0 (0.0) | 1 (2.6) | 22 (56.4) |  |
| **2. It would be easy for you to get information about genetics if you wanted to.** |  |  |  |  |
| **Pre-test N (%)** | **Post-test N (%)** |  |  | **McNemar's test p-value\*** |
|  | Disagree | Neutral | Agree | 0.80 |
| Disagree | 2 (5.4) | 0 (0.0) | 1 (2.7) |  |
| Neutral | 1 (2.7) | 2 (5.4) | 1 (2.7) |  |
| Agree | 1 (2.7) | 1 (2.7) | 28 (75.7) |  |
| **3. You would be able to understand information about how genes can affect your health.** |  |  |  |  |
| **Pre-test N (%)** | **Post-test N (%)** |  |  | **McNemar's test p-value\*** |
|  | Disagree | Neutral | Agree | 0.11 |
| Disagree | 1 (2.6) | 1 (2.6) | 0 (0.0) |  |
| Neutral | 0 (0.0) | 4 (10.3) | 0 (0.0) |  |
| Agree | 1 (2.6) | 4 (10.3) | 28 (71.8) |  |
| **4. You have a good idea about how genetics may influence risk for disease generally.** |  |  |  |  |
| **Pre-test N (%)** | **Post-test N (%)** |  |  | **McNemar's test p-value\*** |
|  | Disagree | Neutral | Agree | 0.11 |
| Disagree | 3 (9.1) | 0 (0.0) | 1 (3.0) |  |
| Neutral | 2 (6.1) | 4 (12.1) | 3 (9.1) |  |
| Agree | 0 (0.0) | 0 (0.0) | 20 (60.6) |  |
| **5. You have a good idea about how your own genetic makeup might affect your risk for disease.** |  |  |  |  |
| **Pre-test N (%)** | **Post-test N (%)** |  |  | **McNemar's test p-value\*** |
|  | Disagree | Neutral | Agree | 0.95 |
| Disagree | 3 (7.9) | 2 (5.3) | 0 (0.0) |  |
| Neutral | 1 (2.6) | 5 (13.2) | 5 (13.2) |  |
| Agree | 0 (0.0) | 5 (13.2) | 17 (44.7) |  |
| **6. You would be able to explain to others how genes affect health.** |  |  |  |  |
| **Pre-test N (%)** | **Post-test N (%)** |  |  | **McNemar's test p-value\*** |
|  | Disagree | Neutral | Agree | 0.04 |
| Disagree | 5 (12.8) | 6 (15.4) | 1 (2.6) |  |
| Neutral | 2 (5.1) | 6 (15.4) | 6 (15.4) |  |
| Agree | 2 (5.1) | 0 (0.0) | 11 (28.2) |  |

For objective knowledge, patients had a mean “genetic knowledge test” score of 0.72 (range: 0-1) in the pre-test (**Table 3**). “Genetic knowledge test” score was significantly different by income group (ANOVA p-value= 0.001) and race (t-test p-value= 0.04) yet did not differ significantly by age (ANOVA p-value= 0.55), education (ANOVA p-value= 0.15), or insurance (ANOVA p-value= 0.06). Patients in the highest income group had significantly higher mean scores, and white women had significantly higher mean test scores than non-white women, while patients with private insurance had a non-significant numerically higher score. There was no significant change in score from pre-to post-test (Wilcoxon p-value= 0.57; **Figure 3**). “Genetic knowledge” was not significantly correlated with any of the four validated psychological measures (data not shown). We assessed correlations between CES-D, BAI, CASE, TPS, “genetic knowledge,” and number of FM recommended therapies in the post-test survey. Patients had a mean of 13.3 (range: 1-36) recommended therapies. We found no significant correlations between number of therapies with CES-D, BAI, CASE, TPS, or “genetic knowledge”.

**Table 3.** Objective Knowledge about Genetics

|  |  |  |
| --- | --- | --- |
| **1. It is possible to see a gene with the naked eye. (False).** |  |  |
| Pre-test % correct<br>(N=58) | Post-test % correct<br>(N=40) | McNemar's test p-value<br>(N=40) |
| 77.2 | 87.2 | **0.04** |
| **2. Healthy parents can have a child with a hereditary disease. (True)** |  |  |
| Pre-test % correct<br>(N=58) | Post-test % correct<br>(N=40) | McNemar's test p-value<br>(N=40) |
| 92.7 | 92.3 | 0.32 |
| **3. The carrier of a disease gene may be completely healthy. (True)** |  |  |
| Pre-test % correct<br>(N=58) | Post-test % correct<br>(N=40) | McNemar's test p-value<br>(N=40) |
| 91.2 | 87.2 | >0.99 |
| **4. Genes are inside cells. (True)** |  |  |
| Pre-test % correct<br>(N=58) | Post-test % correct<br>(N=40) | McNemar's test p-value<br>(N=40) |
| 60.7 | 59.0 | >0.99 |
| **5. A gene is a piece of DNA. (True)** |  |  |
| Pre-test % correct<br>(N=58) | Post-test % correct<br>(N=40) | McNemar's test p-value<br>(N=40) |
| 70.2 | 61.5 | 0.48 |
| **6. All body parts have all of the same genes. (True)** |  |  |
| Pre-test % correct<br>(N=58) | Post-test % correct<br>(N=40) | McNemar's test p-value<br>(N=40) |
| 33.3 | 28.2 | 0.56 |
| **7. A person's race and ethnicity can affect how likely they are to get a disease. (True)** |  |  |
| Pre-test % correct<br>(N=58) | Post-test % correct<br>(N=40) | McNemar's test p-value<br>(N=40) |
| 80.7 | 89.7 | **0.03** |

**Figure 3.**
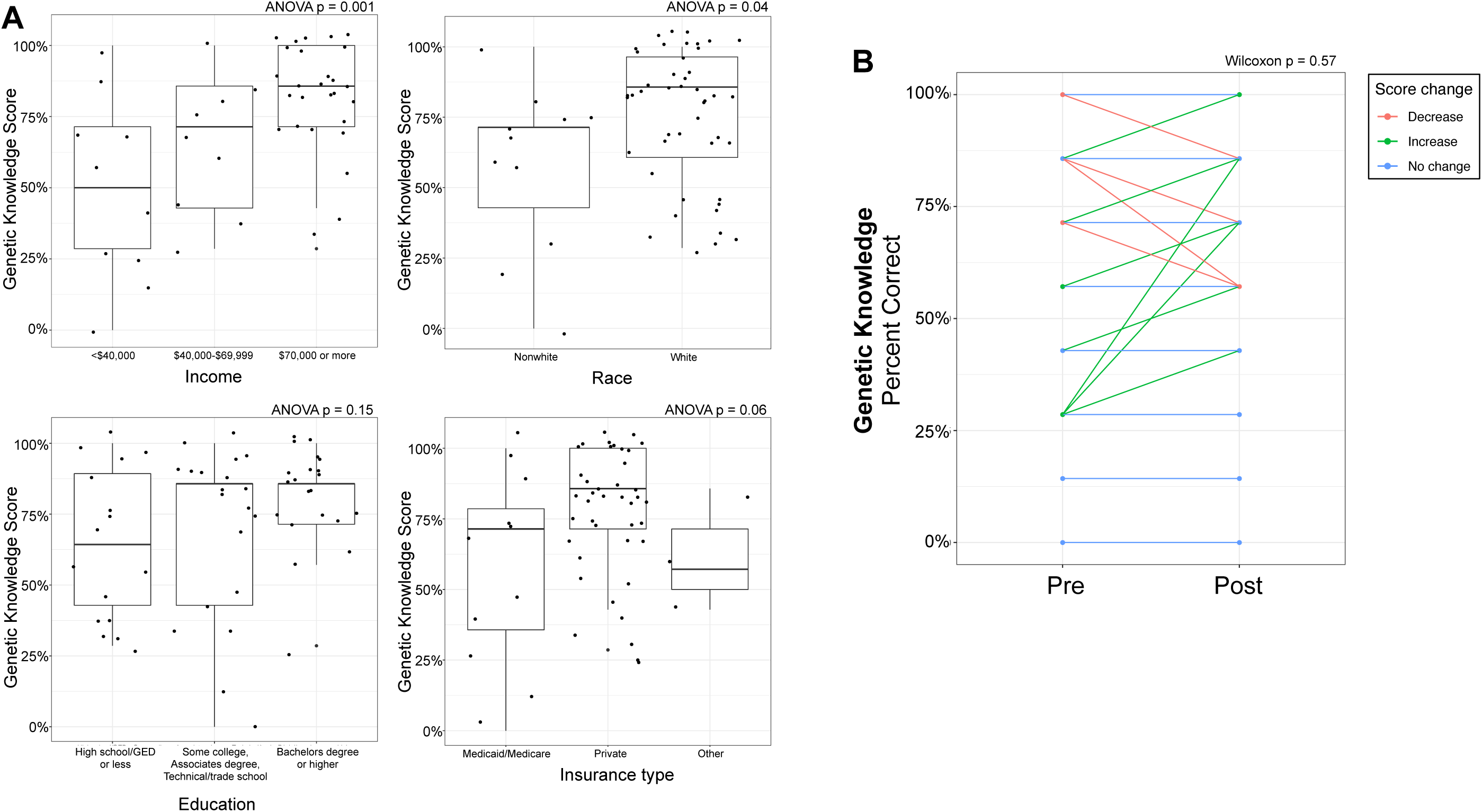
Patient Objective Genetic Knowledge Assessment. Participants completed a seven-question objective genetic knowledge survey at study entry and ‘genetic knowledge score’ assessed as percentage correct. **A**. Association of genetic knowledge score with demographic features including income (top left panel), race (top right), education (bottom left), and insurance type (bottom right). Test of association by ANOVA test indicated with p-value. **B**. Change in genetic knowledge score from study entry (‘Pre-test’) to end of study (‘Post-test). Association was assessed by Wilcoxon signed rank test.

### Patient Motivation, Perception, and Information Seeking Behavior

We administered 9 questions to evaluate patient motivations and perceived risks and benefits of participation in the study (**Supplementary Table 2)**. 32.7% (18/55) of patients believed that finding out their cancer had a high chance of progressing would be too much to handle emotionally. However, most patients (85.7%; 48/56) did not agree that information about their cancer was best left unknown. Most patients were not concerned (58.9%; 33/56) about FM testing being new. Furthermore, most patients (77.4%; 41/53) did not believe they would lose their job if the genomic testing results got out. About half of patients (56.4%; 31/55) believed the FM results would help them change behaviors to reduce disease risk. Most patients also believed (63.6%; 35/55) that the results would help them to seek medical attention to reduce disease risk. To assess information seeking behavior, we administered one question asking patients what steps they have taken to learn more after learning their study results. Most patients (85%) sought information after their genomic testing. 57.5% of patients asked their physician or other provider for information and 27.5% of patients used the internet, their physician, and/or other sources.

## DISCUSSION

As somatic genomic testing becomes increasingly adopted as part of standard care for MBC, there is a critical need to assess its impact on patients outside of implications for clinical care. In this study, patients received identical genomic testing (Foundation Medicine) with results delivered immediately after progression providing a consistent setting for evaluation of patient understanding and perception of genomic testing.

There is growing awareness of differences in genetic testing comprehension, as has been documented previously in the scope of germline genomic testing and race,^28^ and in this study patient understanding of somatic genomic testing showed substantial variability. We used a non-validated 7-question metric to assess genetic knowledge and this demonstrated a wide range of baseline understanding of genetics, with a range 0% to 100% of questions answered correctly in both pre- and post-test. This suggests that providers cannot assume any baseline knowledge of genetics when offering somatic genomic tumor testing. In terms of sociodemographic patient factors, participants who identified as non-white and participants with lower annual incomes had significantly lower baseline genetic comprehension, however, the limited sample size prevented evaluation of possible confounders, such as literacy, numeracy, and other metrics of socioeconomic status. It is important to note that education was not associated with baseline genetic knowledge, possibly skewed by the fact that the observed cohort was predominantly well-educated, limiting our ability to detect an association.

We hypothesized that patients’ genetic knowledge may improve over their time on a prospective clinical trial of somatic genomic testing. There was no formal educational intervention as part of this study but during the informed consent process patients received information on the nature of genetics and genetic testing, the information it could yield, and how that information could impact care. However, our data do not demonstrate any significant change in genetic comprehension from pre-to post-study. While objective genetic comprehension remained stable, patients’ self-assessed ability to explain genetics’ relationship with health to others increased. This suggests a potentially inflated self-appraised ability and understanding, an area explored in the perception of patients’ health and in students’ assessment of knowledge, but not before in the context of patient health education to our knowledge.^29^ There is evidence that information seeking can falsely increase a patient’s subjective knowledge within the domain of genetic testing and we did see that most patients (85%) sought information after their genomic testing.^30^ It is important for physicians to be aware of this phenomenon because it may influence patients concealing health illiteracy.^31^ Collectively, these findings reinforce the need for interventions to educate patients about their care or research studies and to target health literacy, particularly in the informed consent process for genetic/genomic testing.

To investigate patient understanding of genomic testing, we evaluated exploratory questions regarding patient motivations, expectations, perceived risks, and perceived benefits. Notably, the majority of patients mistakenly believed that the NGS test results would explain how they could change their own behavior to lower their risk of disease. This demonstration of advanced cancer patients’ unrealistic expectations of somatic genomic testing corroborates and further informs recent study findings that indicated over 90% of patients believed the NGS test would tell them if they had a higher risk for noncancerous diseases (e.g. Alzheimer’s Disease) or would test for viruses like Human Immunodeficiency Virus despite the informed consent process explicitly stating these would not be accounted for.^32^

In cancer care, there has been a growing focus on mental health, because of the preliminary documentation of associations between depression and worse survival outcomes in women with advanced breast cancer ^16^. Although still disputed, it has been hypothesized that psychological distress, including anxiety and depression, may elicit disease progression by interfering with the hypothalamic pituitary adrenal axis.^33^ While further study is warranted to explore the relationship between psychological health and clinical outcomes, we began to explore the impact of tumor genomic testing on emotional wellbeing. Our cohort of participants had a slightly higher average CES-D depression score (13.6) at baseline in comparison to another trial evaluating depressive-symptoms alone in MBC, which had an average CES-D score of 11.1.^16^ A recent meta-analysis assessed depression in patients with breast cancer worldwide and found that depression was prevalent in 32.2% of patients, similar to our study population with a prevalence of 38% with a CES-D score reflective of depression. In terms of anxiety, 47% of women displayed anxiety in our cohort.^34^ In comparison to other advanced cancer types, such as patients with late stage ovarian cancer with a mean BAI score of 16.88-19 at baseline, our cohort had less severe anxiety on average (mean score of 11.3). However, our study cohort had a slightly greater proportion of patients who experienced at least mild anxiety (47%) in comparison to patients with late stage ovarian cancer (44%).^35^ Interestingly, anxiety and depression positively correlated in our study, indicating that psychosocial interventions for MBC should be comprehensive and not only focus on depression <u>or</u> anxiety. Patients’ mental health status remained rather stable over the duration of the study.

This is the first study to our knowledge to evaluate MBC patients’ trust in their doctor using the validated TPS metric. Patients’ trust in their medical oncologist remained stable over the duration of the study with an average TPS score of 48.4-49.4 (pre- and post-survey, respectively). This is higher than other documented TPS study scores in rheumatic disease and primary care patients which ranged between 41.9 and 45.7, respectively.^36,37^ The stability of the TPS score was interesting given our prior finding that patients lost confidence in their cancer treatment success after undergoing the genomic testing.^20^

Self-efficacy in cancer care, defined as patients’ confidence in maneuvering through their cancer care and relationship with their medical oncologist, trended downward from before to after genomic testing. One possible explanation of this finding is that the physicians unintentionally overstated the ability of the NGS test to impact treatment and patients subsequently became disappointed with the results. Within the context of CASE-cancer, this suggests that patients lose confidence in their perceived capacity to navigate their cancer care from before to after the NGS testing. Because increased self-efficacy, amongst other patient psychological factors and communication styles with their physician, is correlated with better disease outcomes, future study of ways to preserve patient self-efficacy is needed.^38^ One potential method to preserve patient self-efficacy is through increased patient education and empowering supportive interventions.^38-40^

As genomic testing becomes increasingly integrated into clinical care, it is important to evaluate how the translation of this valuable information holistically affects patients. While other studies are beginning to evaluate patient comprehension and perceptions of genomic testing in other disease groups,^41^ this exploration in cancer care has primarily focused on germline genetic testing.^42-44^ These studies still support the need for greater attention on the translation of genomic testing into clinical care, as similar concerns about patient misconceptions about results and what the tests offer are apparent.^41^

Our study does have limitations. There was a relatively limited sample size, partly due to a lower response rate of 58% (58/100). The study still offers unique insight into critical areas regarding the translation of somatic genomic testing into clinical care because of its longitudinal design and utilization of multiple metrics. It is important to note that our use of non-validated metrics to evaluate patient comprehension of genetics, subjective perception of genetic knowledge, or patient perceived risks and benefits of genetic testing does limit our study. Further, the single-center design also supports the need for further exploration of these areas.

In conclusion, this is the first study to longitudinally evaluate the role of depression, anxiety, trust in physician, self-efficacy, objective genetic knowledge, subjective assessment of genetic knowledge, motivations and expectations in regards to somatic genomic testing in a single advanced cancer population. Further exploration of these areas in a larger study are warranted and will hopefully inform how to best support patients who are undergoing tumor genomic testing by better explaining the role of NGS in cancer care through the creation of patient-centered education initiatives.

## Data Availability

Data available upon request.

## Conflict of Interest Statement

### Ethical Approval and consent to participate

All procedures performed in studies involving human participants were in accordance with the ethical standards of the institutional and/or national research committee and with the 1964 Helsinki declaration and its later amendments or comparable ethical standards.

### Consent for publication

All authors agree with publication.

### Informed Consent

All patients completed informed consent approved by the Ohio State University Institutional Review Board, which included a description of the somatic genomic testing and discussion of study risks and benefits with their treating physician.

### Author’s contributions

All authors contributed to data collection, interpretation, and manuscript preparation.

## Acknowledgements

The authors would like to acknowledge the clinical support of Catherine Carson, Celia Garr, Krysten Brown, and Katherine Tyson.

## Supplementary Data

**Supplementary Table 1.**
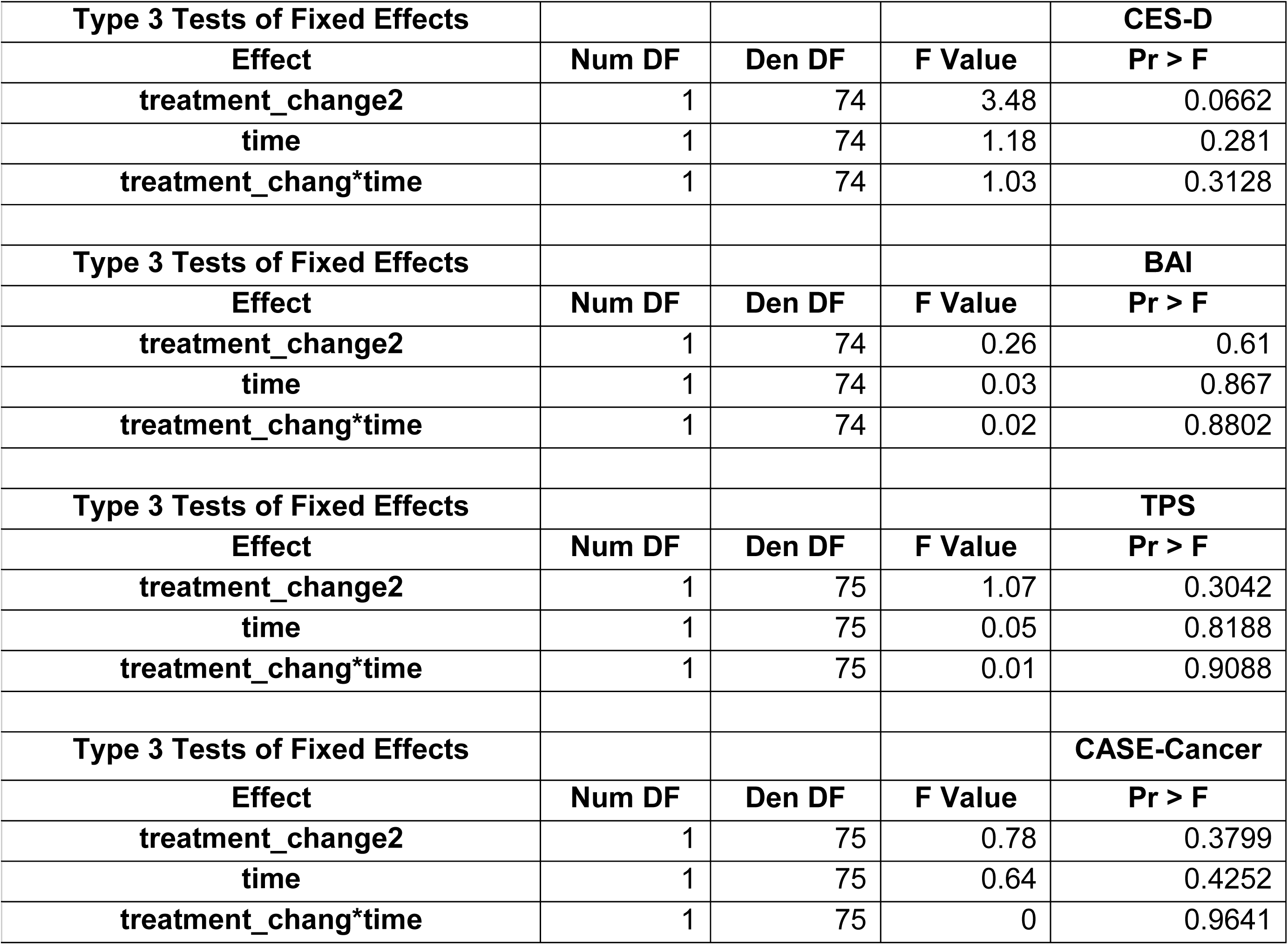
Mixed Effects Model

**Supplementary Table 2.** Patient Motivations and Perceived Risks/Benefits

| *Patient Motivations and Perceived Risks/Benefits*<br>(Pre-test results, N=58*) |  |  |
| --- | --- | --- |
| **Finding out that my cancer has a high chance of progressing would be more than I could handle emotionally.** |  |  |
| Disagree N(%) | Neutral N(%) | Agree N(%) |
| 26 (47.3%) | 11 (20.0%) | 18 (32.7%) |
| **This information about one's cancer is better left unknown.** |  |  |
| Disagree N(%) | Neutral N(%) | Agree N(%) |
| 48 (85.7%) | 6 (10.7%) | 2 (3.6%) |
| **I am concerned about the test because it is new and hasn't been used widely.** |  |  |
| Disagree N(%) | Neutral N(%) | Agree N(%) |
| 33 (58.9%) | 16 (28.6%) | 7 (12.5%) |
| **I am concerned that the test being so new prevents me from asking other patients about their experiences with it.** |  |  |
| Disagree N(%) | Neutral N(%) | Agree N(%) |
| 32 (58.2%) | 16 (29.1%) | 7 (12.7%) |
| **The results will help me change my behaviors and reduce my disease risk.** |  |  |
| Disagree N(%) | Neutral N(%) | Agree N(%) |
| 5 (9.1%) | 19 (34.6%) | 31 (56.4%) |
| **The results will help me seek medical attention and reduce my disease risk.** |  |  |
| Disagree N(%) | Neutral N(%) | Agree N(%) |
| 2 (3.6%) | 18 (32.7%) | 35 (63.6%) |
| **I could lose my job if the results get out.** |  |  |
| Disagree N(%) | Neutral N(%) | Agree N(%) |
| 41 (77.4%) | 10 (18.9%) | 2 (3.8%) |
| **I may learn that I have an increased risk for a disease that I did not want to know about.** |  |  |
| Disagree N(%) | Neutral N(%) | Agree N(%) |
| 30 (53.6%) | 12 (21.4%) | 14 (25.0%) |
| **I may learn that I have an increased risk for a disease that I can do nothing about.** |  |  |
| Disagree N(%) | Neutral N(%) | Agree N(%) |
| 20 (35.7%) | 14 (25.0%) | 22 (39.3%) |

\*N per question ranges from 53-56 due to skipped questions

**Supplementary Figure 1.**
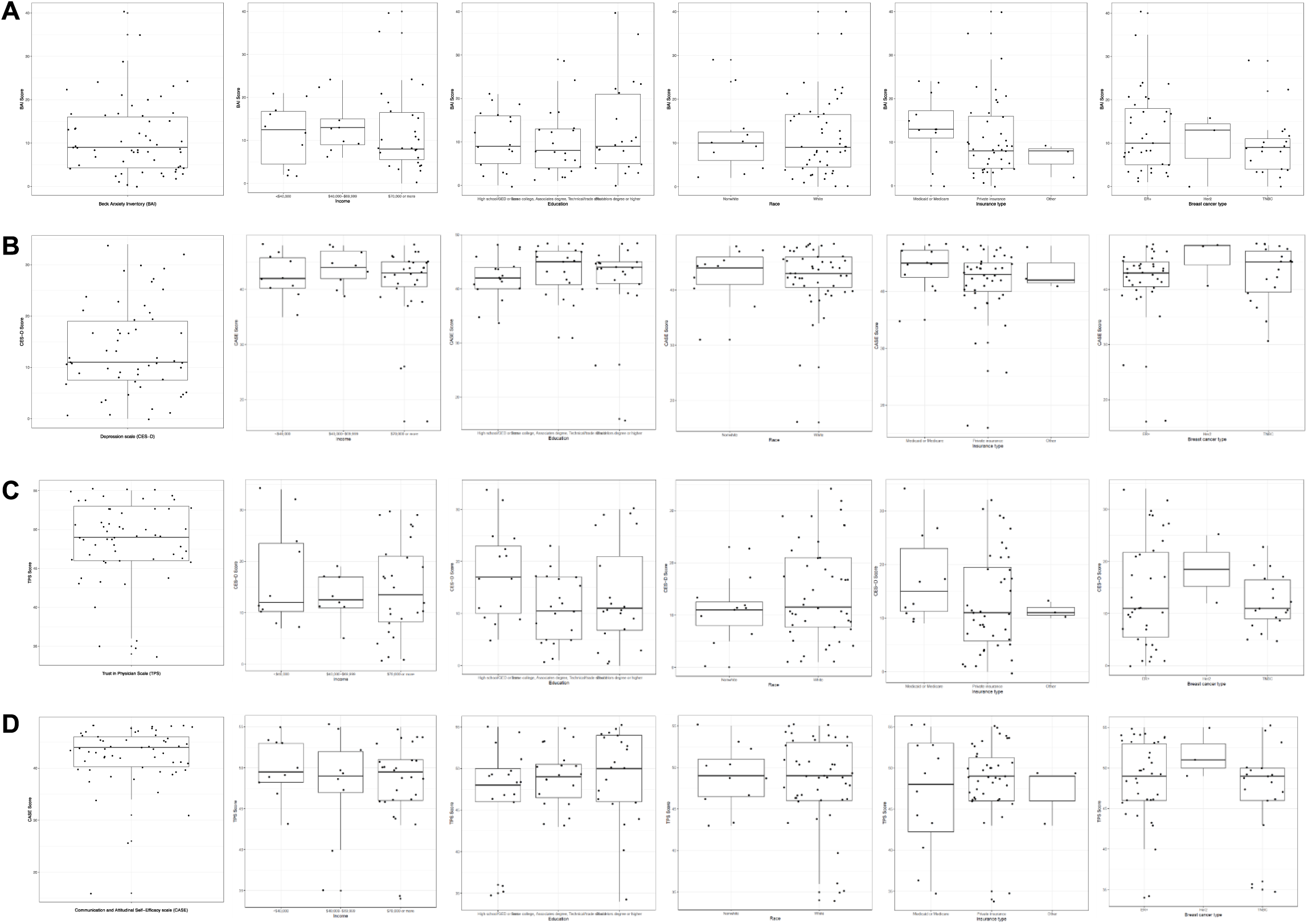
Association of Validated Depression, Anxiety, Physician Trust, and Self-Efficacy Measures with Patient Demographic Features. Four validated psychosocial measures were assessed in patients at study entry: Center for Epidemiologic Studies Depression Scale (**A;** CES-D)[23], Beck Anxiety Inventory (**B;** BAI) [24], Trust in Physicians/Providers Scale (**C;** TPS)[25], and Communication and Attitudinal Self-Efficacy scale for cancer (**D;** CASE-cancer)[26]. There were no significant differences in any of the validated metrics by income (all ANOVA p>0.05), education (all ANOVA p>0.05), race (all t-test p>0.05), insurance (all ANOVA p>0.05), or breast cancer type (all ANOVA p>0.05).

